# Delays, masks, the elderly, and schools: first COVID-19 wave in the Czech Republic

**DOI:** 10.1101/2020.11.06.20227330

**Authors:** Jan Smyčka, René Levínský, Eva Hromádková, Michal Šoltés, Josef Šlerka, Vít Tuček, Jan Trnka, Martin Šmíd, Milan Zajíček, Tomáš Diviák, Roman Neruda, Petra Vidnerová, Luděk Berec

**Affiliations:** Center for Theoretical Studies, Husova 4, 11000 Praha 1, Czech Republic; CERGE-EI, Politických vězňů 7, 11121 Praha 1, Czech Republic; New Media Studies, Faculty of Arts, Charles University, Na Příkopě 29, 11000 Praha 1, Czech Republic; Department of Mathematics, University of Zagreb, Bijenička 30, 10000 Zagreb, Croatia; Department of Biochemistry, Cell and Molecular Biology, Third Faculty of Medicine, Charles University, Ruská 87, 100 00 Praha 10, Czech Republic; Czech Academy of Sciences, Institute of Information Theory and Automation, Pod Vodárenskou věží 4, 18200 Praha 8, Czech Republic; Department of Criminology, School of Social Sciences, University of Manchester, Oxford Rd, Manchester, UK; Czech Academy of Sciences, Institute of Computer Science, Pod Vodárenskou věží 2, 18200 Praha 8, Czech Republic; Centre for Mathematical Biology, Institute of Mathematics, Faculty of Science, University of South Bohemia, Branišovská 1760, 37005 České Budějovice, Czech Republic; Czech Academy of Sciences, Biology Centre, Institute of Entomology, Department of Ecology, Branišovská 31, 37005 České Budějovice, Czech Republic

## Abstract

Running across the globe for more than a year, the COVID-19 pandemic keeps demonstrating its strength. Despite a lot of understanding, uncertainty regarding the efficiency of interventions still persists. We developed an age-structured epidemic model parameterized with sociological data for the Czech Republic and found that (1) delaying the spring 2020 lockdown by four days produced twice as many confirmed cases by the end of the lockdown period, (2) personal protective measures such as face masks appear more effective than just a reduction of social contacts, (3) only sheltering the elderly is by no means effective, and (4) leaving schools open is a risky strategy. Despite the onset of vaccination, an evidence-based choice and timing of non-pharmaceutical interventions still remains the most important weapon against the COVID-19 pandemic.

**One sentence summary:** We address several issues regarding COVID-19 interventions that still elicit controversy and pursue ignorance

---

The COVID-19 pandemic of the SARS-CoV-2 virus has held the world in its grip for more than a year now, with many countries still facing severe epidemic waves (*1*). With the first three cases reported on March 1, 2020, the epidemic in the Czech Republic (Czechia) started and was initially fueled by Czech citizens returning from the alpine ski resorts of Italy and Austria. Population-wide interventions began on March 11, 2020, with the closing of schools, soon followed by travel restrictions, closing of restaurants, sports and cultural facilities and shops (with some exceptions), as well as the introduction of a duty to wear face masks and keep at least 2 m inter-personal distance in public (Table 1). By May 2020 the epidemic situation in Czechia stabilized and the lockdown restrictions were gradually relaxed. During the summer months only sporadic local outbreaks occurred and life returned nearly to normal. All started to change once again by the end of August 2020, resulting in an unprecedented second and third waves, accompanied by a worrying increase in the numbers of hospitalized individuals, including those requiring intensive care (*2, 3*).

**Table 1:** Population-wide interventions against COVID-19 (lockdown) in the Czech Republic during the spring first wave.

| Date | Intervention |
| --- | --- |
| March 11, 2020 | Schools closed, home office recommended where possible |
| March 12, 2020 | Travelling restricted |
| March 14, 2020 | Closing of restaurants, sports and cultural facilities, and shops (with exceptions) |
| March 19, 2020 | Duty to wear face masks, keep at least 2 m inter-personal distance, and use disinfection on public |

Since lockdowns not only damage national economy but also negatively affect the mental state of people (*4*), governments all over the world tend not to implement severe population-wide restrictions until they become unavoidable. Fortunately, we can learn from past epidemic waves and increase the effectiveness of any forthcoming interventions. How fast do we need to react in order to limit potential cases and casualties? How can we protect ourselves effectively or limit social contacts when wearing hazmat suits or locking ourselves at home for two or three weeks are not viable options? Can we limit the worst impacts of the epidemic just by sheltering the elderly with the rest living more or less normally? What about leaving schools open or not forcing people to work from home? These questions still sprout controversies among politicians and in the general public not only in Czechia, and appear to have not been explored deeply in the scientific literature.

Here we develop a mathematical model of the COVID-19 epidemic to address these four questions. The model is structured by age, type of inter-individual contacts (at home, school, work and in the community), and space, and considers all important epidemic classes (see Materials and Methods in Supplementary materials). We parameterize the model by combining the public health data on the spring 2020 first wave in Czechia, including the type and timing of interventions implemented, sociological data from surveys carried out before and during this wave, real-time population movement data, and data published in the scientific literature. In order to account for all these sources of information and their inevitable uncertainty we used the Approximate Bayesian Computation (ABC) framework based on massive super-computing simulations. This technique, used to estimate parameters of complex models in genomics and other biological disciplines (*5, 6*), including epidemiology (*7–9*), allows us to assess credibility intervals of our predictions (Materials and Methods).

## Using initial phase and lockdown period as baseline scenario

Our baseline scenario, to which we compare a number of alternatives, concerns the initial phase of the epidemic and the subsequent lockdown, i.e. the period from the start of the epidemic up to May 7, 2020, to which all interventions outlined in Table 1 applied. As the starting date we arbitrarily used February 1, 2020, in order to cover the real and unknown beginning of the epidemic (Methods; the first three cases apparently became infected and showed symptoms before March 1 and some undetected asymptomatic cases were most likely imported too). The mechanistic character of our model allows for an exact implementation of specific dates of initiation of various interventions and for the setting up of factors that reflect the impacts of these interventions on epidemiological as well as behavioral parameters. We base these factors on data from sociological surveys behavior before and during the lockdown (Methods, Fig. S1).

As the first step in applying the ABC technique we simulated 100,000 realizations of epidemic dynamics with parameter values generated randomly from prior distributions based on public health data in Czechia and on literature-based data (Fig. 1A; Materials and Methods). The 0.1% of model realizations (100) that best matched the observed age-specific cumulative numbers of confirmed cases were then selected using a rejection-sampling algorithm (Fig. 1B-D). The parameter sets corresponding to the selected realizations form an outer estimate of a posterior distribution of parameter values, the distribution that possesses residual uncertainty in the parameters after the model is confronted with the observed data (*5–7*) (Fig. 1A). One may think of the selected parameter sets as representing different worlds, the observations of which are compatible with the actual observations, and study the epidemic of COVID-19 in any of these worlds or in all of them simultaneously (Methods). We believe our model is applicable to dynamics beyond the COVID-19 epidemic in Czechia (Fig. 1).

**Fig. 1:**
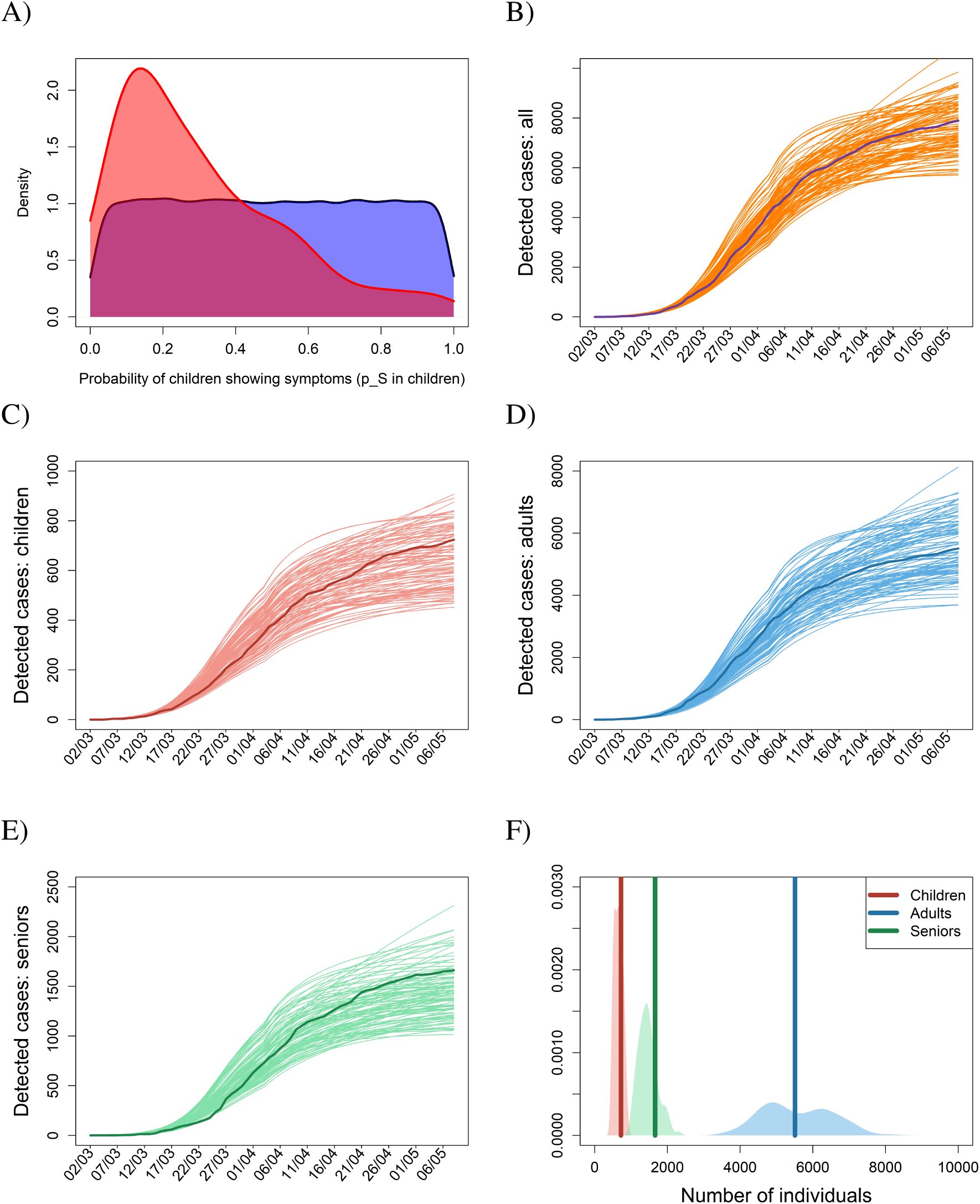
Approximate Bayesian Computation (ABC). (A) Prior parameter distributions based on available information are first built for every estimated parameter (blue). (B) Parameter sets randomly generated from these priors are used to run the model; the sets for which the model outcome provides good fit to the observations are selected and used to built a posterior distribution over the estimated parameters (red). (C-F) Temporal dynamics of the age-specific cumulative numbers of confirmed cases for the baseline scenario, covering the initial phase of epidemic and the following lockdown, for (C) children (age cohort 0-19 years), (D) adults (age cohort 20-64 years), and (C) elderly (age cohort 65+ years), with their sum plotted as panel B. (F) Probability density functions for the numbers of confirmed cases by 7 May 2020. In panels B-E, dark curves represent real observations, while the light (orange in B) curves are results of model simulations run for 100 selected posterior parameter sets. In panel F, the dark vertical lines represent the observed numbers of confirmed individuals by 7 May 2020, while the light areas are density plots for the simulated numbers of confirmed individuals by 7 May 2020 for the 100 selected posterior parameter sets.

## Impact of timing and potential alternative interventions

Several scenarios alternative to the actually implemented lockdown and motivated by ideas appearing among the general public are considered, run for the selected parameter sets, and compared with the baseline scenario.

The first two scenarios address one of the most important issues in containing an incipient epidemic: the timing of interventions. We find that establishing every single intervention four days earlier (scenario R1) or later (scenario R2) than actually happened produces significant differences in the numbers of confirmed cases by May 7, 2020 (Fig. 2). All else being equal, any delay of four days in implementing the lockdown thus doubled the number of confirmed cases by May 7, 2020. When deciding on interventions and implementing them time is undoubtedly of essence.

**Fig. 2:**
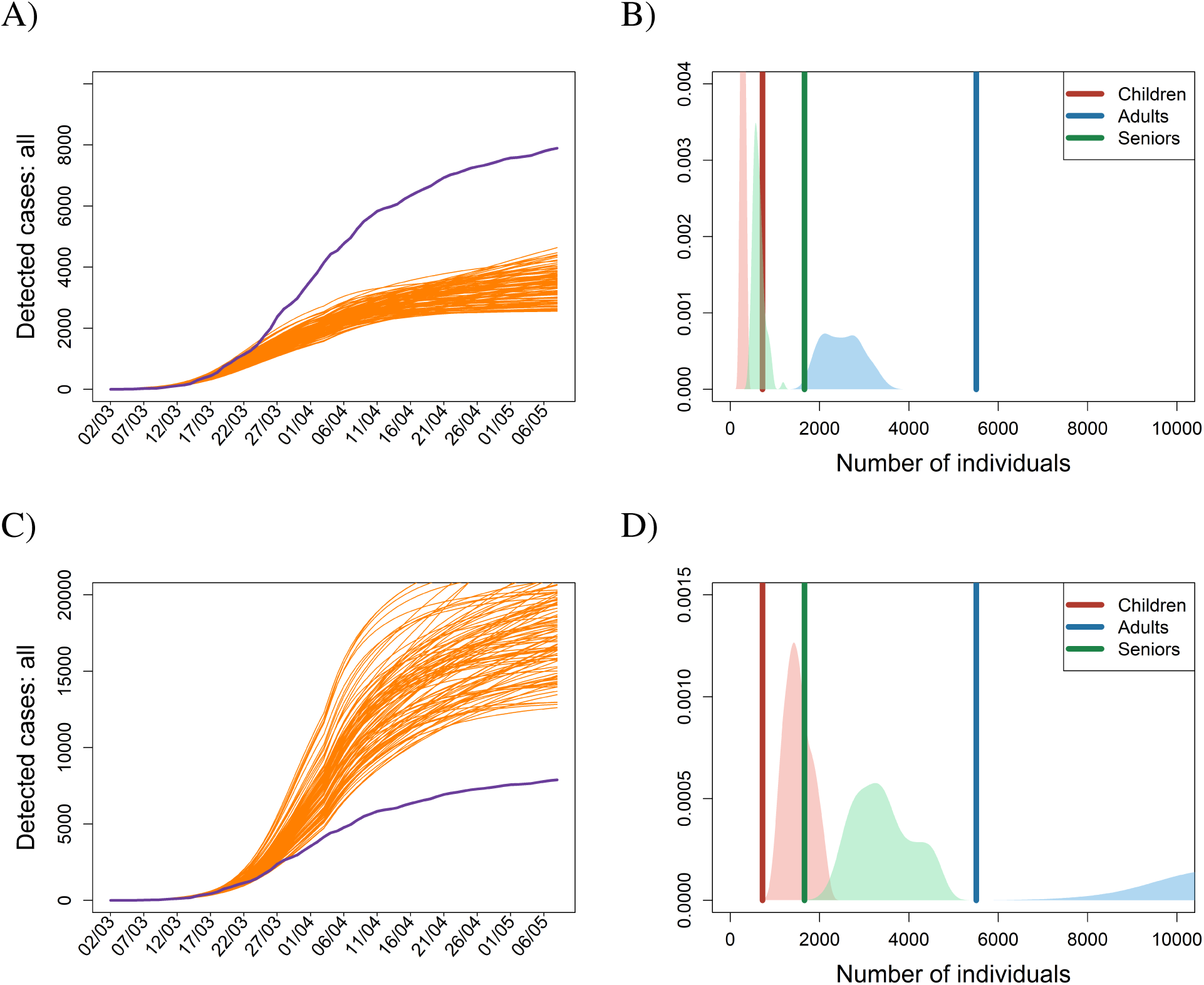
Lockdown scenario shifted in time. Temporal dynamics of the cumulative numbers of confirmed cases for the lockdown scenario shifted four days backward (A-B) or four days forward (C-D). Blue lines in the left panels represent real observations of the total cumulative numbers, while the orange lines are the results of model simulations run for the 100 selected posterior parameter sets. In the right panel, probability density functions for the age-specific numbers of confirmed cases by 7 May 2020 are provided; the dark vertical lines represent the observed numbers of confirmed individuals by 7 May 2020, while the light areas are density plots for the simulated numbers of confirmed individuals by 7 May 2020 for the 100 selected posterior parameter sets.

Our next two scenarios are motivated by the initial reluctance to order the compulsory wearing of face masks in many countries, and by the existence of groups that deny their utility even today. However, personal protection is not only about face masks. It also comprises increased hygiene and a behavioral change towards protecting oneself. Setting the amount of interpersonal contacts to the sociologically observed level while neglecting any sort of personal protection, produced twenty times more confirmed cases by May 7, 2020 (scenario R3, Fig. 3A and B). On the contrary, sticking to the level of personal protection observed in the population and not limiting interpersonal contacts worsened the epidemic approximately by a factor of five by May 7, 2020 (scenario R4, Fig. 3C and D). The modeled epidemic thus appears less sensitive to changes in contact structure than to changes in the factors reducing infection transmission due to personal protective measures.

**Fig. 3:**
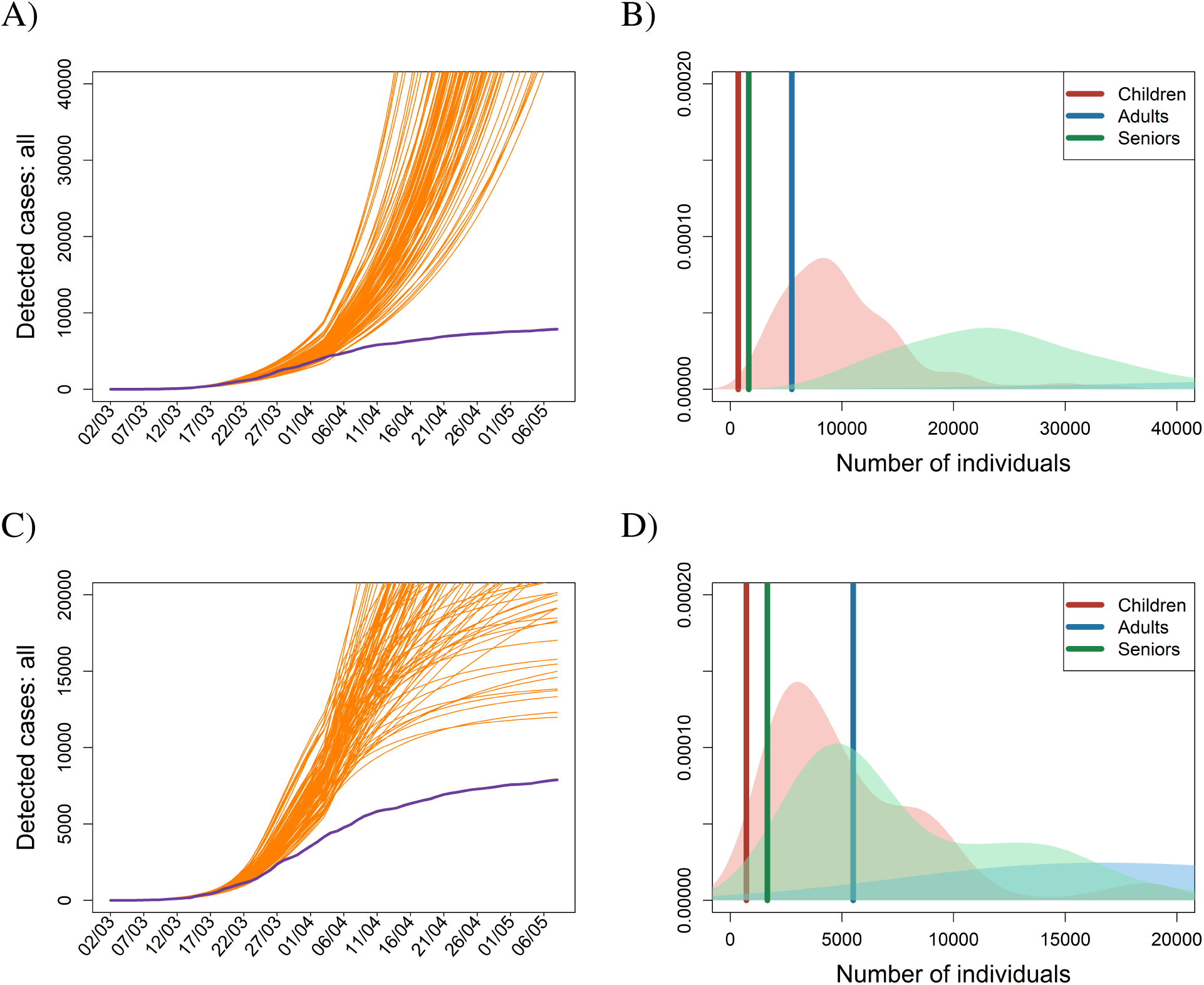
Limiting contacts versus enhancing protection. Dynamics of the cumulative numbers of confirmed cases for (A-B) the scenario R3 with reducing compliance to use personal protective measures to zero and (C-D) the scenario R4 without limiting contact structure. Legend as in Fig. 2.

A strategy of sheltering the elderly while letting the remaining population mix relatively freely remains a popular mantra among opinion makers dissenting to population-wide restrictions. Unfortunately, such a strategy does not appear to work (Fig. 4). The explanation for this is, perhaps surprisingly, quite straightforward. The elderly typically represent a relatively small but still sizeable fraction of the population (about 20% of people in Czechia are 65 years or older). It is thus impossible to isolate them completely and the extraordinarily high viral load that would occur in the freely mixing younger population would eventually bring the infection even into the protected group. In either of the two scenarios modeling this situation (R5 and R6) the number of affected seniors roughly doubles or triples by May 7, 2020 compared to baseline but even more alarming is the convex rather than concave shape of the curve indicating a continuing rapid increase after that date (Fig. 4).

**Fig. 4:**
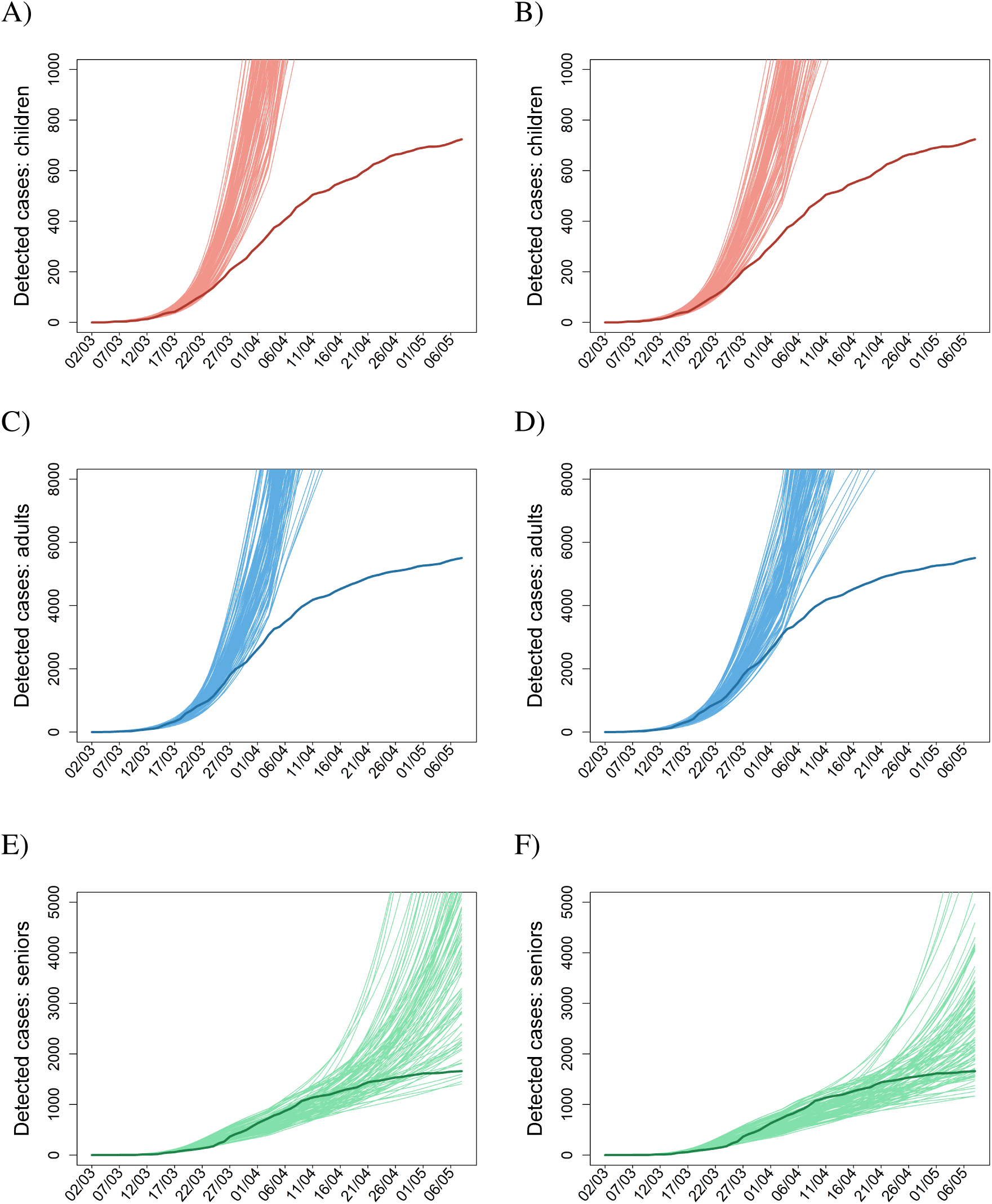
Sheltering the elderly does not work. (A-C) Contacts of seniors with the rest of population are reduced by 90%, while contacts among the rest of the population are reduced by 10%. (D-F) Contacts of seniors with the rest of population are reduced by 90%, while contacts among the rest of the population are reduced by 30%. In both scenarios, personal protection in seniors set to 90%, while in children and adults only to 50%. Legend as in Fig. 1.

Finally, an ongoing debate in many countries is whether to leave schools open (or rather whether to open them, under some restrictions or rotation schemes). In a similar vein one may ask whether home office or limiting contacts at work is effective. In our model leaving schools open means on average 4.8 more child-child contacts per day, while no home office similarly keeps on average up to 5.3 more adult-adult contacts per day (*10*). Leaving all other restrictions in place, the last two scenarios (R7 and R8) demonstrate the importance of these restrictions (Fig. 5A and C). With open schools or no home office the total number of confirmed cases by May 7, 2020 increases compared to baseline. Since these interventions predominantly affect different parts of the population (children vs. adults), we observe differential effects on the respective age cohorts (Fig. 5B and D). Yet the more interesting observation here is the uncertainty in the qualitative course of infection not observed in any of the previously considered scenarios: whereas some trajectories level off as for the baseline scenario, suggesting a small effect of the respective interventions, others continue to rise (Fig. 5).

**Fig. 5:**
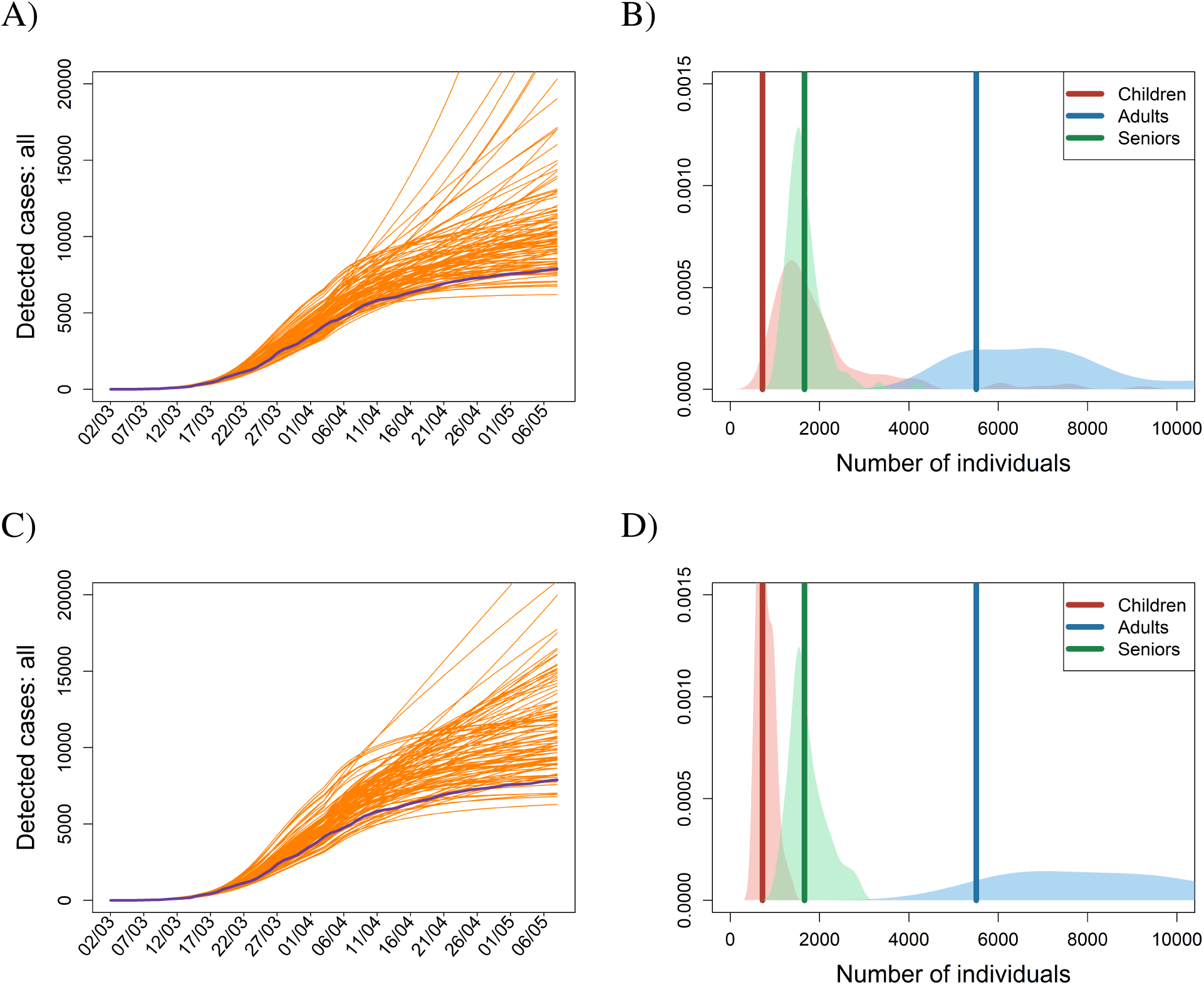
Letting schools or work fully open. Dynamics of the cumulative numbers of confirmed cases for (A-B) the scenario R7 with schools left open and (C-D) the scenario R8 with not enforcing any home office. Legend as in Fig. 2.

A way of assessing the effect of (not) implementing an intervention in our multi-world framework is to calculate the impacts of an intervention relative to the baseline scenario when both simulations come from the same parametric world. This way of plotting the results showcases our findings that every four days of delaying the interventions result in a doubling of the number of confirmed cases by the end of the lockdown period, that personal protective measures are more effective than just reducing the number of contacts, that sheltering the elderly is not as effective as it may seem, and that leaving schools open or not adopting home office has ambiguous effects (Fig. 6). It also shows that despite different parameter sets that nonetheless produce results matching actual observations, the effects of interventions are comparable across these parametric worlds, providing a robustness check for our model-based assessment of their efficacy.

**Fig. 6:**
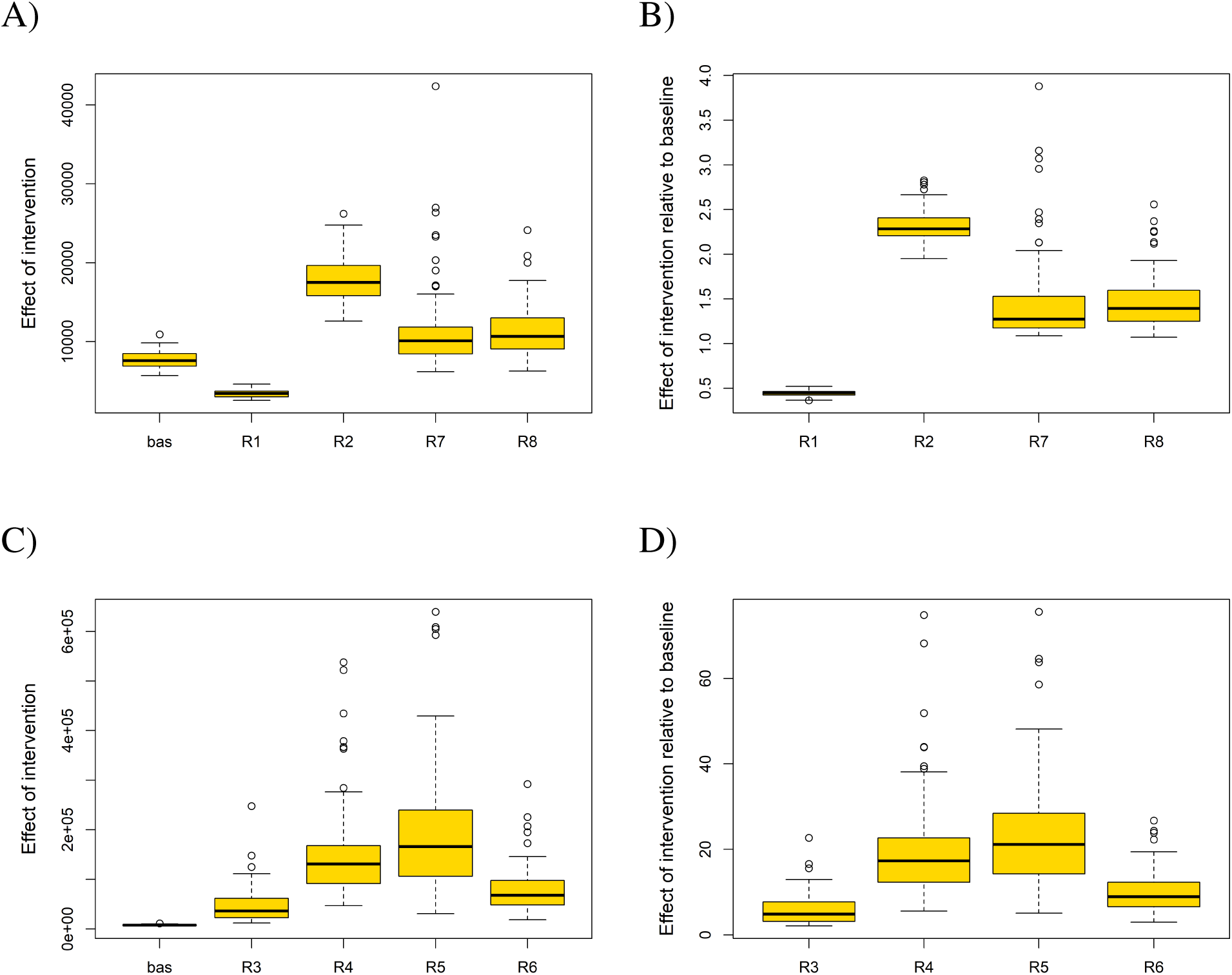
Comparing effects of analyzed scenarios. Box plots showing the effects of adopted scenarios in absolute terms (A-B) and relative to the baseline general lockdown scenario (C-D) for the total numbers of confirmed cases expected on 7 May 2020. Codes for different scenarios are described earlier in the text.

## Discussion

Non-pharmaceutical interventions always underlie the first wave of defence against an incipient epidemic. The art of responding to the epidemic is not only in setting up the right restrictions and in the right time but also in ordering them optimally and implementing them as effectively as possible. This all sounds intuitive, and it really is. However, governments still vary in their approaches.

The best scenarios from a theoretical point of view, such as timely and adequate testing and tracing and full tracing coverage, are unfortunately seldom achieved in practice. Mathematical models demonstrate their strength in situations when the optimal is unattainable, showing, for example, what we can expect and how to respond to testing delays of one or two days or when only 70-80% tracing coverage is doable (*11*). The major advantage of modeling epidemics is thus in providing quantitative assessments of what kind and/or intensity of interventions is likely (not) enough to prevent serious epidemic impacts, or, more loosely, which strategies of containing epidemic would (not) work.

Two types of models have mostly been used to provide insights into effects of non-pharmaceutical interventions in controlling COVID-19 epidemics. One type, exemplified by (*12–16*), is a mechanistic prospective model that aims at predicting a future course of epidemic given a set of alternative scenarios. Such models may be calibrated on real observations in a specific country, and typically provide a relative ranking of the considered strategies. Another type of models, exemplified by (*17–21*), are statistical in nature and attempt to reveal effects of interventions by retrospectively analyzing observed time series data on COVID-19 epidemics in many countries at once. Such models typically do not have a means of switching the various interventions on and off, thus losing the ability to explore the impacts of alternative (i.e. not implemented) interaction scenarios. Our model in a sense bridges these two types, providing a mechanistic description of the epidemic with the ability to switch interventions on and off at specific time instants and to modify their intensity while at the same time being calibrated on a robust set of observed data (*22, 23*).

We used a mathematical model to analyze the spring first wave of COVID-19 in Czechia. The model allowed us to distinguish between the effects of personal protective measures and interventions reducing social contacts and to set exact dates of implementing such regulations. It structures the population by age (three cohorts: 0-19 years [children], 20-64 years [adults] and 65 and more years [seniors]), type of contacts (four types: at home, at school, at work and other contacts within community) and space (206 administrative units of the Czech Republic). The model was calibrated on age-specific cumulative numbers of confirmed cases, requiring a testing layer that modeled delays from symptoms onset to testing to reporting and a proportion of the population not willing to test themselves despite being symptomatic. We found that (1) delaying the lockdown by four days led to twice as many confirmed cases by the end of the lockdown period, (2) personal protective measures such as wearing face masks and using hand sanitizers were more effective than only reducing social contacts, (3) sheltering only the elderly while letting the remaining population live relatively normally was not a viable strategy, and (4) leaving schools open and not adopting home office appeared to be risky measures.

A key and unique feature of our model is its use of sociological data quantifying the degree of compliance with the interventions, that is, the degree of interpersonal contact limitation in various environments as well as the degree of compliance with using personal protective measures. Therefore we need not speculate on the degree of compliance in the society but rather work with the actual level of the reduction of social contacts relative to the preceding non-epidemic state. These proportional reductions in contacts enter our model as fundamental system characteristics and serve as benchmarks, towards which we test alternative intervention strategies.

Another unique feature of our modeling approach is how we deal with uncertainty. Although our model is deterministic we calibrate it via a stochastic technique, known as Approximate Bayesian Computation (ABC) (*5–7*), applied to model parameters that are sourced either from the literature or the data sources for the epidemic in Czechia. In this way we generate a number of posterior parameter sets (or different worlds) that all produce outcomes matching the actual observations of the age-specific numbers of confirmed cases, and examine how the epidemic behaves in those worlds when the interventions change. We emphasize that even though our model is calibrated for the epidemic in the Czech Republic it can straightforwardly be recast for any other country or region if relevant data on behavioral and population characteristics are available.

Despite the obvious fact that any delay in implementing interventions aimed at slowing down the epidemic has a negative impact, many politicians and public health officers tend to downplay its importance. Here we quantified this effect for the lockdown applied during the spring first wave of COVID-19 in Czechia, showing that a delay of four days doubles the number of confirmed cases by the end of lockdown period. All else being equal, this also means doubling the number of deaths during that period, as the change affects all age cohorts equally. Clearly, this exact quantitative relationship cannot be expected to hold for any epidemic wave and in any country but it shows that the effect is far from negligible and has to be taken seriously. In light of this result it is quite concerning to see the specifics of lockdown implementation in Czechia: it has been sadly common that interventions announced as a response to a worsening epidemic situation were actually implemented with a delay of one or two weeks. A similar exploration of COVID-19 epidemic in the United States between March 15 and May 3, 2020, suggested that implementing their interventions one or two weeks earlier would have reduced the numbers of confirmed cases and dead by 50% and 90%, respectively (*23*). This further underscores the need to act as quickly as possible.

Any epidemic can only be brought under control by interrupting the chain of transmissions. This can be done by limiting social contacts and/or by adopting personal protective measures. Since locking ourselves at home or wearing full protective suits for two or three weeks are obviously not viable options, it is important to examine the relative effect of limiting contacts versus enhancing protection. In this light it is surprising how long people (and even some governments) resisted using face masks in many parts of the world, with Sweden recommending wearing face masks only in December 2020 (*24*). On the other hand, countries such as Italy or Austria insisted on wearing face masks also during the relatively ‘quiet’ summer months. A positive impact of wearing face masks on limiting the spread of COVID-19 has been demonstrated by a number of modeling studies (*25, 26*). We add to this knowledge our finding that adhering to personal protective measures is likely more effective than just reducing social contacts. It is then not surprising that our simulations also suggested that switching the dates of implementing wearing face masks and closing schools in Table 1 resulted in only half the amount of confirmed cases by May 7, 2020 (results not shown). For any respiratory epidemic to come in the future using face masks and other personal protective measure should therefore be among the first interventions implemented.

The idea that sheltering of elderly, or more broadly any high-risk portion of the population, while letting the rest live more or less normally, is sufficient to prevent deaths still has its followers. We show here that it is deeply flawed. First, in Czechia and many other European countries the age cohort of age 65 years or model forms at about 20% of the population, of which only a small part lives in senior houses while many others live in multi-generational families, so it is difficult to shelter successfully. Second, leaving the rest of the population behaviourally unrestrained would soon result in a high virus prevalence in this group. The virus would then sooner or later percolate into the elderly group despite its relatively high protection. Our simulations show that even a 30% reduction in social contacts among the rest of the population would not help much in preventing the infection of the elderly. Moreover, instead of the decelerating by May 7, 2020, the scenario of sheltering the elderly exhibits a continuing exponential growth. This scenario is thus not a viable option, even more so if we take into consideration the potential impact of such isolation on the mental state of the elderly: (*27*) found that older individuals felt higher depression and greater loneliness during pandemic.

Finally, an issue commonly discussed in both country-specific studies (*22*) and studies spanning more countries (*17, 19*), is whether to leave schools open or to close them. The closing of schools was among the first interventions in many countries (*17, 19*), presumably because of existing pandemic plans tailored to influenza. We considered scenarios in which schools remained open or alternatively home office was absent, with all other interventions kept as during the lockdown, with ambiguous results. Whereas in many parametric worlds an effect of leaving schools open or absenting home office appears negligible, in many remaining worlds an increase in the number of confirmed cases becomes linear rather than decelerating by May 7, 2020. This is worrying since a significant increase in the prevalence among younger cohorts sooner or later manifests itself in the older, inevitably resulting in more deaths. We thus recommend some form of restriction also at schools and at work. Other studies are likewise ambiguous on this issue, and can be generally divided into those that rank closing schools among the most important interventions (*17, and references therein*) and those that claim a relatively small effect of closing schools (*19, 22, 28, 29*). Discordant results of these studies may be attributed to using different type of models, which calls for caution when reading and interpreting such studies. For example, both (*17*) and (*19*) admit that some of their conclusions might have been affected by the fact that the analyzed interventions were applied in a specific order or close to each other. Lockdown is an expensive measure and should generally be applied only when milder interventions fail. With a limited knowledge of COVID-19 and the resulting fear during the spring first wave, many countries adopted a form of lockdown, allowing us to take it as a baseline scenario with which to compare alternative strategies and thus help assess interventions for the other epidemic waves. We used a well-defined model that allowed us to play with the adopted interventions and at the same time calibrate the model on the lockdown situation. Our results suggest that a combination of a timely application of personal protective measures, the limiting of contacts and an effective testing and contact tracing constitute an optimal strategy. This may sound trivial but mathematical models can tell us how much is not enough, what does not work and what appears to be a necessary minimum to keep the society alive and to protect its health.

## Data Availability

Data used in the manuscript were either taken from the literature or provided by state institutions of the Czech Republic. In the latter case, they are not publicly avilable, but are acknowledged in the manuscript. The software code used to run the model is available (without publicly unavailable data) on reasonable request.

## Acknowledgments

The authors thank The Institute of Health Information and Statistics of the Czech Republic (IHIS CR) for providing us detailed data on many characteristics of the COVID-19 epidemic in Czechia. We also acknowledge the PAQ Reseach agency and the Median agency for sharing with us the results from their sociological surveys. The mobility data were provided by the project “RODOS Transport Systems Development Centre” which is supported by the Technology Agency of the Czech Republic. This paper uses data from SHARE Waves 1, 2, 3, 4, 5, 6 and 7 (DOIs: 10.6103/SHARE.w1.700, 10.6103/SHARE.w2.700, 10.6103/SHARE.w3.700, 10.6103/SHARE.w4.700, 10.6103/SHARE.w5.700, 10.6103/SHARE.w6.700, 10.6103/SHARE.w7.700), see (*30*) for methodological details.

## Funding

The SHARE data collection has been funded by the European Commission through FP5 (QLK6-CT-2001-00360), FP6 (SHARE-I3: RII-CT-2006-062193, COMPARE: CIT5-CT-2005-028857, SHARELIFE: CIT4-CT-2006-028812), FP7 (SHARE-PREP: GA No 211909, SHARE-LEAP: GA No 227822, SHARE M4: GA No 261982) and Horizon 2020 (SHARE-DEV3: GA No 676536, SERISS: GA No 654221), and by DG Employment, Social Affairs & Inclusion. Additional funding from the German Ministry of Education and Research, the Max Planck Society for the Advancement of Science, the U.S. National Institute on Aging (U01 AG09740-13S2, P01 AG005842, P01 AG08291, P30 AG12815, R21 AG025169, Y1-AG-4553-01, IAG BSR06-11, OGHA 04-064, HHSN271201300071C) and from various national funding sources is gratefully acknowledged (see www.share-project.org). This work was supported by the Ministry of Education, Youth and Sports of the Czech Republic through the project SHARE-CZ+ (CZ.02.1.01/0.0/0.0/16 013/0001740). VT was supported by the QuantiXLie Center of Excellence, a project co-financed by the Croatian Government and European Union through the European Regional Development Fund - the Competitiveness and Cohesion Operational Programme (KK.01.1.1.01.0004).

## Author Contributions

LB, JS and RL conceived the study. RN, PV, MŠ, EH and MŠ consulted the design and development of the model. JŠ, JT, VT and MZ acquired, managed and analyzed data on COVID-19 epidemic in Czechia. LB and JS coded the model and performed the simulations. All authors wrote and revised the manuscript.

## Author Competing Interests

All authors declare to have no competing interests

## Data and materials availability

Model code allowing to reproduce all simulations is available from the authors upon request (will be made available in a code storage place when the manuscript is accepted).

## Supplementary materials

Materials and Methods

Supplementary Text

Fig. S1

Tables S1 to S3

References *(31-38)*

## Materials and Methods

Our epidemic model is an extension of the classic SEIR model, structured by age, type of contacts and space. We start with describing an unstructured version of the model. Its extension to age, various types of contacts and space then follows.

### Core model

Due to contacts with infectious individuals, susceptible individuals (*S*) may become exposed (*E*), that is, infected but not yet infectious (the process of infection transmission is described below). The exposed individuals then become either asymptomatic for the whole course of infection (*I*_*a*_, with a probability 1 *− p*_*S*_) or presymptomatic for just a short period of time before becoming symptomatic (*I*_*p*_, with a probability *p*_*S*_). The *I*_*p*_ individuals later become symptomatic. We assume that a proportion *p*_*T*_ of such symptomatic individuals decide to undergo testing for the presence of SARS-CoV-2 (*I*_*s*_). On the contrary, the proportion 1 *− p*_*T*_ of symptomatic individuals (most likely those with very mild symptoms) decide not to undergo testing and stay at home (*I*_*h*_). Since in the Czech Republic, deaths attributed to COVID-19 did not generally occur outside hospitals, the *I*_*a*_ and *I*_*h*_ individuals eventually recover (*R*). Deaths outside hospitals were an important issue in countries heavily hit by the COVID-19 spring wave where hospital capacities were soon depleted (Italy, Spain).

Considering discrete time, with one time step corresponding to one day, our model so far consists of the following equations

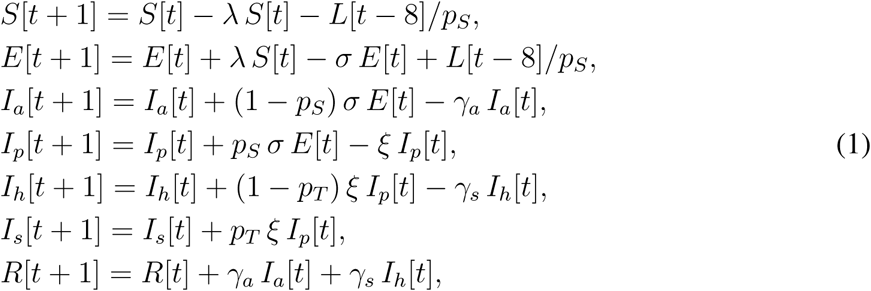

where the parameters *σ, ξ, γ*_*a*_, and *γ*_*s*_ represent probabilities with which individuals leave the respective model classes. These probabilities are related to the mean duration an individual spends in each such class. All model variables are summarized in Table S1, and all model parameters in Tables S2 and S3.

The force of infection *λ* in the model (1) sums contributions from all infectious classes, that is, *I*_*a*_, *I*_*p*_, *I*_*h*_, and *I*_*s*_:

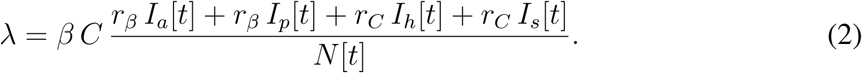

Here, *β* is a probability of infection transmission upon contact between susceptible and infectious individuals, *C* is the contact rate (the mean number of other individuals an individual has an effective contact with per day), *r*_*β*_ is a factor reducing the infection transmission probability for an asymptomatic individual relative to a symptomatic one, *r*_*C*_ is a factor reducing the contact rate of a symptomatic individual relative to an asymptomatic one (having symptoms should force an individual to reduce contacts with others), and *N*[*t*] is the total population size at time *t*. We emphasize here that the calculation of *λ* will be modified further in the text, as we extend our model description.

### Initial state

The hitherto unexplained variable *L*[*t*] accounts for the imported COVID-19 cases from abroad, mostly from Italy and Austria. A list of all confirmed (symptomatic) imported cases is available at onemocneni-aktualne.mzcr.cz/covid-19. However, we do not introduce such imported cases as symptomatic. Rather, we assume they came earlier as exposed, and introduce them before they were actually tested positive (to account for a delay between exposition and confirmation). Moreover, to account for the likely situation that some of the imported cases remained undetected as being asymptomatic for the whole course of infection, we divide the number of known imported cased by *p*_*S*_, the probability of exposed individuals eventually becoming symptomatic.

### Testing layer

This layer links the number of symptomatic individuals to the number of confirmed cases. The period from the onset of symptoms, through sampling and subsequent processing, up to infection confirmation and case isolation is assumed to take *d*_*T*_ days (testing delay). Symptomatic individuals that undergo testing are assumed to be always tested positive (we do not assume false negatives). We thus redefine equation for *I*_*s*_ as:

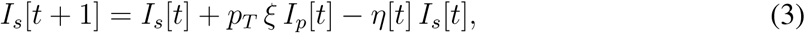

where *η* = 1 *−* exp(*−*1*/d*_*T*_) is the testing rate. The testing rate *η*[*t*] commonly increases in time, as the whole testing process becomes more efficient as the epidemic unfolds. We use data on the COVID-19 epidemic in Czechia to quantify the testing delay separately for March and for April and May (Table S2).

The number of newly (positively) tested symptomatic individuals at day *t* thus equals *η*[*t*] *I*_*s*_[*t*]. Therefore, the total number of such individuals yet to be reported (*B*) is

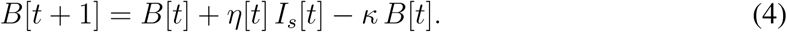

Here *κ* is the publication rate, calculated as *κ* = 1 *−* exp(*−*1*/d*_*P*_), where *d*_*P*_ is the period from case confirmation to case reporting (publication delay). Although this rate may change in time, too, it was relatively constant during the spring first wave in Czechia. The total number of confirmed cases reported until and including time *t* + 1 (*K*) is therefore

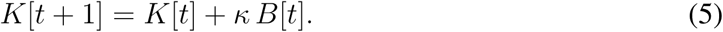

### Dynamics in hospitals

A proportion *p*_*H*_ of (positively) tested symptomatic individuals (those with relatively severe symptoms) require hospitalization, whereas the remaining proportion 1 *− p*_*H*_ (those that have only mild symptoms) are sent home to isolation (*I*_*z*_) until recovery. Hospitalized individuals may follow several pathways, depending on the number of hospital states one considers. Whereas most published models did not consider any class of hospitalized individuals (*14*), the others considered one to three hospital states: one to cover all hospitalized individuals (*31*), two to distinguish between common hospital beds and ICUs (*12*), and three to further detach ICU patients that need lung ventilators or ECMOs (*32*). We consider here one hospital state, for which we introduce three parameters: the proportion *p*_*D*_ of hospitalized individuals that eventually die, mean duration from hospital admission to death *d*_*HD*_, and mean duration from hospital admission to recovery *d*_*HR*_. We thus introduce two hospital classes, *H*_*D*_ and *H*_*R*_, representing the hospitalized individuals that eventually die or recover, respectively, and extend the above model equations as follows:

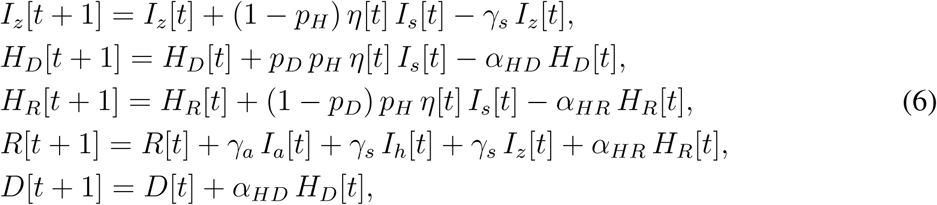

where *α*_*HX*_ = 1 *−* exp(*−*1*/d*_*HX*_) for *X* = *D, R*.

A modification is also required for calculation of the force of infection *λ*. In the formula (2),

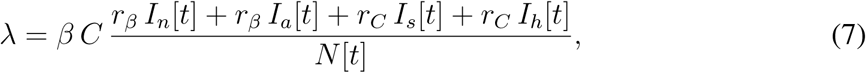

the denominator *N* [*t*] needs to be reinterpreted as the total population size at time *t* except those individuals that have already died (*D*), are isolated at home (*I*_*z*_) or are hospitalized (*H*_*D*_, *H*_*R*_). Individuals in these four classes are expected not to transmit infection to others.

### Age structure

As SARS-CoV-2 is known to differently impact children, adults and seniors (*22*), we distinguish three major age classes: 0-19 years (children), 20-64 years (adults), and 65+ years (seniors). These classes interact via the force of infection. Both the probability of infection transmission upon contact *β* and the daily number of contacts *C* are now 3 *×* 3 matrices, referred to below as the transmission matrix and the contact matrix, respectively.

The transmission matrix *β* is assumed to have the following structure:

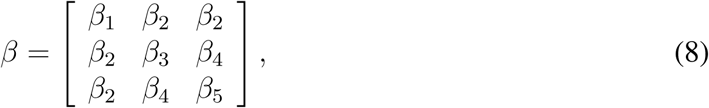

where *β*_1_ is a transmission probability between two children, *β*_2_ is a transmission probability between children and adults or seniors, *β*_3_ is a transmission probability between two adults, *β*_4_ is a transmission probability between seniors and adults, and *β*_5_ is a transmission probability between seniors. We estimate parameters *β*_1_, *β*_2_, *β*_3_, *β*_4_, and *β*_5_ by fitting our model to data on the age-specific cumulative numbers of confirmed cases.

The contact matrix *C* describes the mean number of other individuals of any age cohort (rows) that an individual of an age cohort (columns) meets per day; (*10*) published such a contact matrix for 152 countries, including the Czech Republic. Moreover, they expressed it as a sum of four specific contact matrices describing daily numbers of contacts at home (*C*_*H*_), school (*C*_*S*_), work (*C*_*W*_), and of other types of contact (*C*_*O*_):

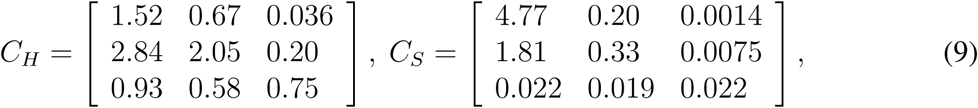

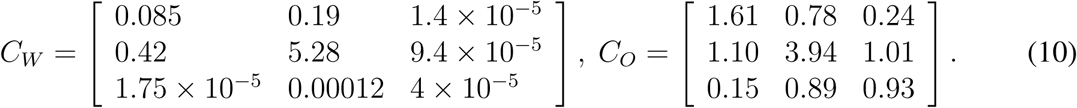

We exploit this division when defining and exploring various realistic intervention strategies.

Once infected, individuals of each age cohort proceed independently of individuals of other age cohorts. However, some model parameters that decide on specific pathways through the model are made age-specific. These are probabilities of becoming symptomatic *p*_*S*_, undergoing testing *p*_*T*_, requiring hospitalization *p*_*H*_, and dying *p*_*D*_ (Tables S2 and S3). Last but not least, since we know age of any imported COVID-19 case in the initial phase of epidemic, we assign each such case to the appropriate age cohort.

### Spatial structure

Spreading of any pathogen in a population, including current epidemic of SARS-CoV-2, has a spatial component. To model this process, we split the Czech Republic geographically into well-defined 206 administrative units. For each such unit, the population size and its proportional distribution into the three considered age classes is known (Czech Statistical Office, www.czso.cz/csu/czso/home). Moreover, we compose and use a 206 *×* 206 mobility matrix that gives mean daily mobility patterns of individuals between all pairs of the spatial units (proportions of individuals travelling per day from a unit to another). Only individuals from the classes *S, E, I*_*a*_, *I*_*p*_, and *R* are allowed to travel in our model. At each time step (one day), our age-structured model is first run (independently) in each spatial unit, and then the mobility matrix is applied to the updated populations in each unit. Regarding the imported cases in the initial phase of epidemic, we have information about the broader region in the Czech Republic each case comes from, so we assign a random spatial unit from the corresponding region to each such case.

The mobility matrix was constructed by averaging mobility patterns obtained from a telecommunication company across two weeks. Two such matrices were used, one representing the normal state using data from January 2020, and the other representing the lockdown state using data from the second half of March 2020. Both matrices were recalculated to account for the company’s market share. Furthermore, movement matrices were adjusted to account for average visiting time of 6 hours, i.e. all movement intensities were divided by 4. Due to the lack of age-specific data, these matrices were assumed identical for all three age classes.

### Sociological data

Our baseline scenario considers all interventions that were in effect during the lockdown established in March 2020 (Table 1). In modeling those interventions, we exploit a division of the contact matrix *C* into four matrices describing contacts at home (*C*_*H*_), school (*C*_*S*_), work (*C*_*W*_), and other types of contacts (*C*_*O*_). Starting from the corresponding dates listed in Table 1, we multiply the respective matrices by factors *r*_*H*_ = 0.44 for home, *r*_*W*_ = 0.45 for work, *r*_*S*_ = 0.03 for schools, and *r*_*O*_ = 0.35 for other types of contacts. Moreover, personal protection, activated on 19 March 2020, including wearing face masks on public, wide use of disinfection and keeping inter-individual distances of more than 2 metres on public, was modeled as follows: all elements of the transmission matrix *β* were multiplied by a factor

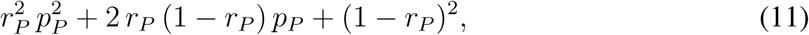

where *r*_*P*_ = 0.88 represents 88% compliance of using personal protection measures (averaging masks and hygiene and computing mean over the high efficiency data; Fig. S1), and *p*_*P*_ is the proportional reduction in transmission due to personal protection estimated via model calibration (Table S3). All these numbers are based on results of public opinion surveys organized by two agencies during lockdown: panel surveys by the PAQ Reseach agency (www.paqresearch.cz) and retrospective questioning by the Median agency (www.median.eu). Results of these surveys, summarized to our modeling purposes, are provided in Fig. S1. These data show that during the second half of March and essentially during the whole April, contacts of all kinds have been largely reduced while personal protection was significant.

### Model calibration

Values of several model parameters remain uncertain, of which the transmission matrix *β* is virtually always one of them. This and some other model parameters, listed in Table S3, are estimated by fitting the simulated cumulative number of confirmed cases in any age class and spatial unit *K*, summed over space but not age, to the age-specific time series on the actually reported cumulative numbers of confirmed cases in the Czech Republic.

There are many ways to meaningfully perform model calibration on real-world data and many optimization and filtering methods exist (*33*). We use the Approximate Bayesian Computation (ABC), a technique used to estimate parameters of complex models in genomics and other biological disciplines (*5–9*). The major advantage of this method is that it naturally works with all sources of uncertainty acknowledged in the model. At the same time, the ABC does not rely on likelihood calculations and in case of sufficient computation power it can be used with models of virtually any complexity. The variant of ABC with rejection sampling that we used

**Figure S1:**
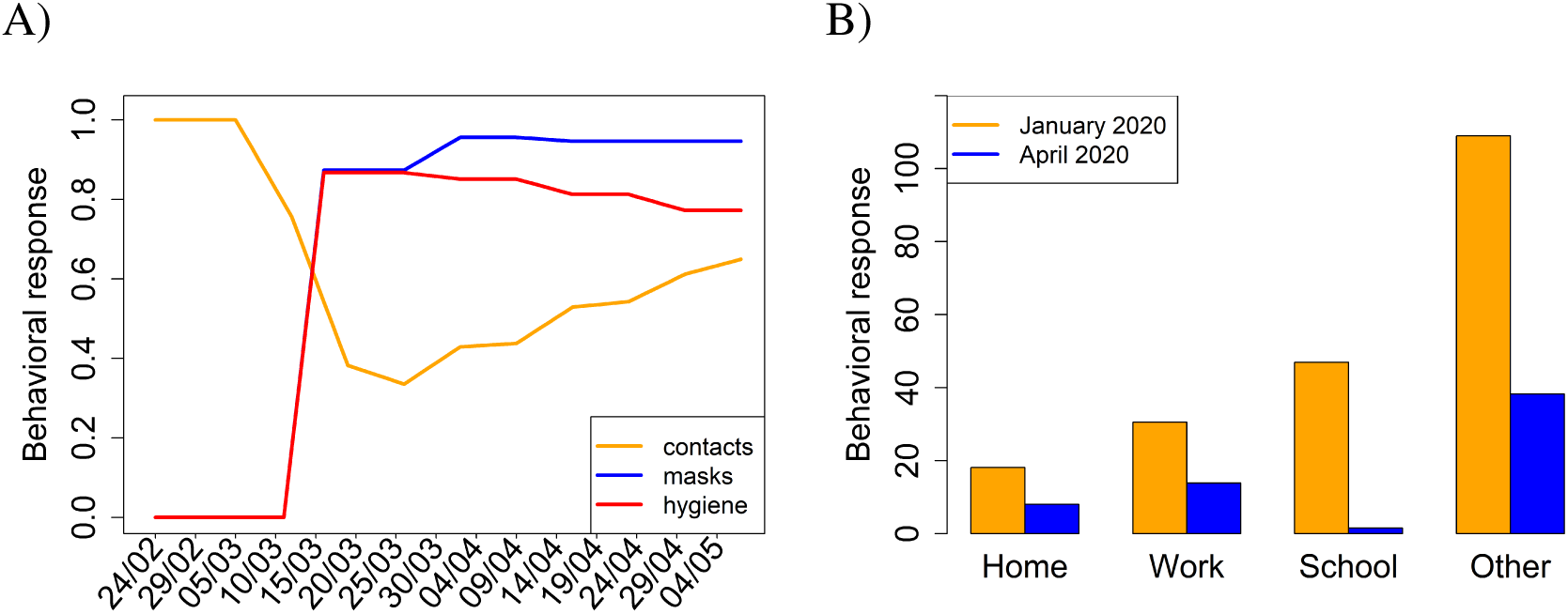
Results of sociological surveys we use in our study. (A) Behavioral responses before and during the lockdown in Czechia, based on panel surveys organized by the PAQ Reseach agency (www.paqresearch.cz) every week (in between the subsequent surveys, data were linearly interpolated). The orange line is a proportional reduction in the numbers of social contacts relative to the pre-pandemic state (value 1), the red and blue lines are proportions of respondents that reported using face masks and increased hygiene. (B) Reduction of the weekly number of contacts in different environments due to the COVID-19 epidemic in Czechia, according to the public opinion polls organized by the Median agency (www.median.eu). The respective proportional reductions in contacts that we used for parameterizing the lockdown period thus are 0.44 for home, 0.45 for work, 0.03 for schools, and 0.35 for other types of contacts; our model distinguishes just these four types of contact.

consisted of three steps. First, we used our model to simulate summary statistics for calibration (the age-specific numbers of confirmed cases; *K*) *N* times (100,000), drawing the uncertain model parameters from prior distributions based on literature and available data on the Czech Republic epidemic; selected prior distributions for the parameters to be estimated are given in Table S3. Second, we compared the simulated summary statistics with the observed one, using the Euclidean distance *D*. Third, we selected model simulations that satisfied *D < ϵ*, where *ϵ* was chosen to pass 0.1% (100) of simulations into the selected set. Given that the used summary statistics are informative, the distribution of parameters corresponding to the selected simulations is known to converge from outside to the Bayesian posterior distribution of parameter values with *ϵ* going to 0, and is referred to as the approximate posterior (*5*). The choice of *ϵ* and *N* in the ABC is driven by compromise between computation power and smoothness and accuracy of the approximate posterior.

The set of selected parametric sets thus allows us to evaluate remaining parameter uncertainty, given the available data and adopted summary statistics (*5–7*). This is crucial to realize, since although different parameter sets may similarly fit the available data (have similar summary statistics), and often provide similar short-term predictions, they may demonstrate significant differences in longer term and in interplay with intervention policies. To cope with such uncertainty, we do not evaluate only an absolute impact of an intervention scenario, but calculate also its impact relative to the baseline scenario, separately for each selected parametric set. If that relative impacts is consistent over whole posterior distribution of parameters, we may have confidence in its potential effect on the epidemic spread. To apply the ABC technique, we use the abc package in R (*34*), modified to work with non-normalized summary statistics.

### Parameterizing individual scenarios

In scenarios R1 and R2, all dates in Table 1 were shifted either four days earlier or four days later. In scenario R3, the level of personal protection *r*_*P*_ was set to 0. In scenario R4, the multiplicative factors *r*_*H*_, *r*_*S*_, *r*_*W*_, *r*_*O*_ for the contact matrices *C*_*H*_, *C*_*S*_, *C*_*W*_, *C*_*O*_ were all set to 1. In scenario R5, we set *r*_*H*_ = *r*_*W*_ = *r*_*S*_ = *r*_*O*_ = 0.9 and *r*_*P*_ = 0.5 for children and adults and *r*_*H*_ = *r*_*W*_ = *r*_*S*_ = *r*_*O*_ = 0.1 and *r*_*P*_ = 0.9 for seniors. In scenario R6, we set *r*_*H*_ = *r*_*W*_ = *r*_*S*_ = *r*_*O*_ = 0.7 and *r*_*P*_ = 0.5 for children and adults and *r*_*H*_ = *r*_*W*_ = *r*_*S*_ = *r*_*O*_ = 0.1 and *r*_*P*_ = 0.9 for seniors. In scenario R7, we kept *r*_*H*_ = 0.44, *r*_*W*_ = 0.45, *r*_*O*_ = 0.35 and set *r*_*S*_ = 1. Finally, in scenario R8, we kept *r*_*H*_ = 0.44, *r*_*S*_ = 0.03, *r*_*O*_ = 0.35 and set *r*_*W*_ = 1.

**Table S1:** List of state and testing-related variables used in the model.

| Variable | Meaning |
| --- | --- |
| $S$ | Susceptible individuals |
| $E$ | Exposed individuals |
| $I_a$ | Asymptomatic individuals for the whole course of infection |
| $I_p$ | Presymptomatic individuals, just before becoming symptomatic |
| $I_s$ | Symptomatic individuals (limit their contacts) |
| $I_h$ | Symptomatic individuals that avoid testing yet also limit their contacts |
| $I_z$ | Positively tested individuals isolated at home |
| $H_R$ | Hospitalized individuals that later recover |
| $H_D$ | Hospitalized individuals that eventually die |
| $R$ | Recovered individuals |
| $D$ | Dead individuals |
| $L$ | Importation cases during the initial epidemic phase |
| $B$ | Positively tested symptomatic individuals to be reported |
| $K$ | Cumulative number of confirmed cases of COVID-19 |

**Table S2:** List of model parameters.

| Parameter | Meaning | Relationship to data |
| --- | --- | --- |
| $\sigma$ | Rate of leaving $E$ class | $1 - \exp(-1/d_E)$ |
| $\xi$ | Rate of leaving $I_p$ class | $1 - \exp(-1/d_P)$ |
| $\gamma_a$ | Rate of recovery from $I_a$ | $1 - \exp(-1/d_A)$ |
| $\gamma_s$ | Rate of recovery from $I_h$ and $I_z$ | $1 - \exp(-d_S)$ |
| $\eta$ | Testing rate | $1 - \exp(-1/d_T)$ |
| $\kappa$ | Publication rate | $1 - \exp(-1/d_P)$ |
| $\alpha_{HX}$ | Rates of leaving hospital states | $1 - \exp(-1/d_{HX}), X = R, D$ |
| Parameter | Meaning | Value |
| $d_T$ | Testing delay* | 6.07 (March), 2.88 (April/May) |
| $d_P$ | Publication delay* | 0.76 |
| $d_A$ | Duration of asymptomatic period | $d_P + d_S$ |
| Parameter | Meaning | Children / Adults / Seniors |
| $p_H$ | Probability of hospitalization* | 0.01 / 0.15 / 0.25 |
| $d_{HR}$ | Time between hospitalization and recovery* | 16.23 / 20.11 / 23.74 |
| $d_{HD}$ | Time between hospitalization and death* | – / 7.60 / 11.31 |
| $p_D$ | Probability of dying when in hospital* | 0 / 0.046 / 0.31 |

**Table S2:**
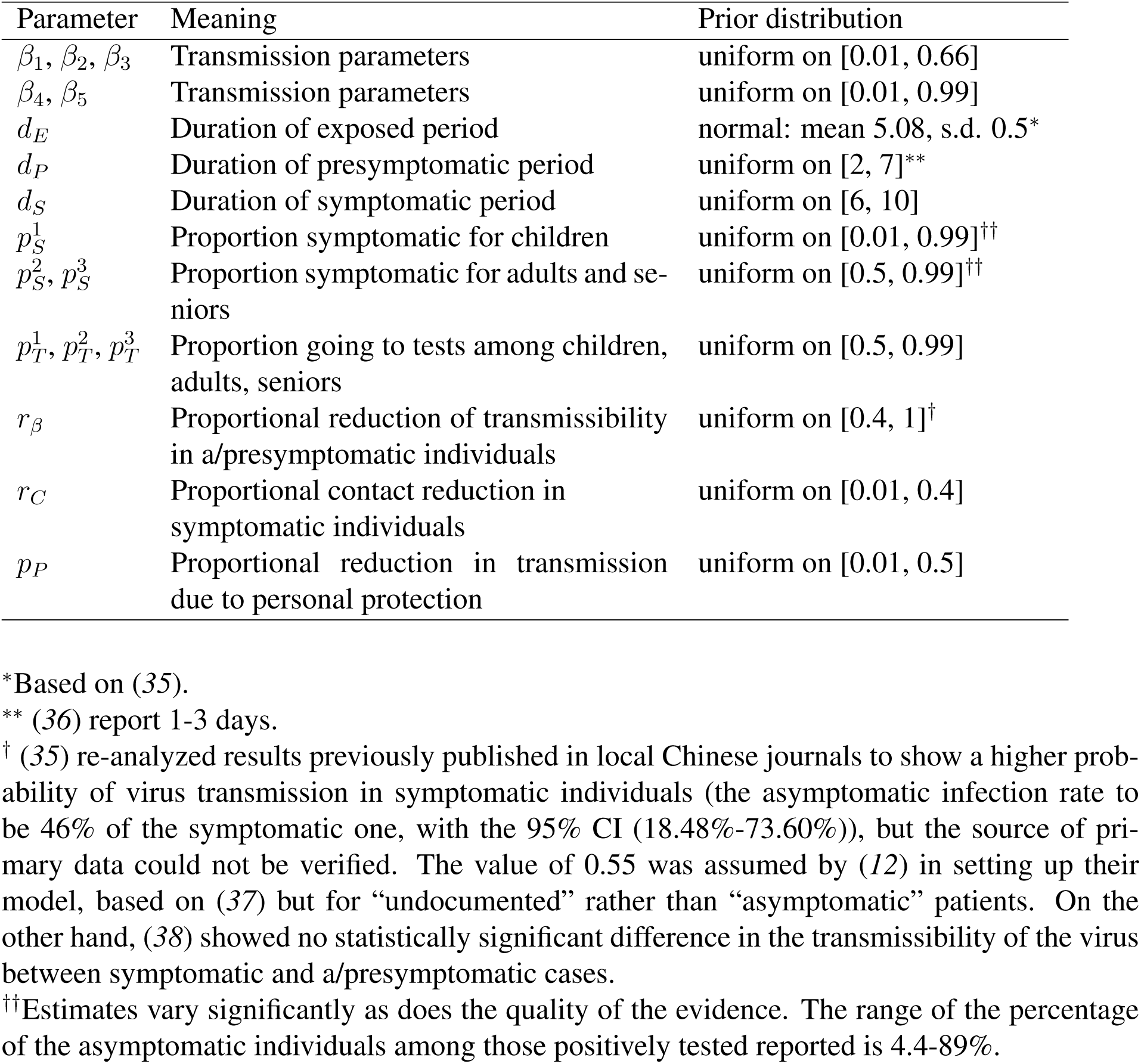
List of parameters estimated by the ABC model calibration technique, including their corresponding prior distributions.

## Notes

### Competing Interest Statement

The authors have declared no competing interest.

### Funding Statement

VT was supported by the QuantiXLie Center of Excellence, a project cofinanced by the Croatian Government and European Union through the European Regional Development Fund - the Competitiveness and Cohesion Operational Programme (KK.01.1.1.01.0004).

### Author Declarations

This is a modelling study that used anonymized data on COVID-19 patients so that no ethics committee approvals are necessary. The use of these data was approved by The Institute of Health Information and Statistics of the Czech Republic.

### Summary of Updates

The manuscript has been revised as a whole, streamlining the text and adding or removing results relative to the previous version to create a more coherent storyline. The idea and methods of the study remain the same as in the previous version.

